# Variation in monitoring: Glucose measurement in the ICU as a case study to preempt spurious correlations

**DOI:** 10.1101/2023.10.12.23296568

**Authors:** Khushboo Teotia, Yueran Jia, Naira Link Woite, Leo Anthony Celi, João Matos, Tristan Struja

**Author notes:** **Corresponding Author** Naira Link Woite, MD, Cell, MIT - Institute for Medical Engineering and Science, Laboratory for Computational Physiology, MIT, E25-530-53 Carleton St, Cambridge, MA 02139. shared first authors (Khushboo Teotia, Yueran Jia). shared last authors (João Matos, Tristan Struja).

## Abstract

**Objective:** Health inequities can be influenced by demographic factors such as race and ethnicity, proficiency in English, and biological sex. Disparities may manifest as differential likelihood of testing which correlates directly with the likelihood of an intervention to address an abnormal finding. Our retrospective observational study evaluated the presence of variation in glucose measurements in the Intensive Care Unit (ICU).

**Methods:** Using the MIMIC-IV database (2008-2019), a single-center, academic referral hospital in Boston (USA), we identified adult patients meeting sepsis-3 criteria. Exclusion criteria were diabetic ketoacidosis, ICU length of stay under 1 day, and unknown race or ethnicity. We performed a logistic regression analysis to assess differential likelihoods of glucose measurements on day 1. A negative binomial regression was fitted to assess the frequency of subsequent glucose readings. Analyses were adjusted for relevant clinical confounders, and performed across three disparity proxy axes: race and ethnicity, sex, and English proficiency.

**Results:** We studied 24,927 patients, of which 19.5% represented racial and ethnic minority groups, 42.4% were female, and 9.8% had limited English proficiency. No significant differences were found for glucose measurement on day 1 in the ICU. This pattern was consistent irrespective of the axis of analysis, i.e. race and ethnicity, sex, or English proficiency. Conversely, subsequent measurement frequency revealed potential disparities. Specifically, males (incidence rate ratio (IRR) 1.06, 95% confidence interval (CI) 1.01 - 1.21), patients who identify themselves as Hispanic (IRR 1.11, 95% CI 1.01 - 1.21), or Black (IRR 1.06, 95% CI 1.01 - 1.12), and patients being English proficient (IRR 1.08, 95% CI 1.01 - 1.15) had higher chances of subsequent glucose readings.

**Conclusion:** We found disparities in ICU glucose measurements among patients with sepsis, albeit the magnitude was small. Variation in disease monitoring is a source of data bias that may lead to spurious correlations when modeling health data.

## INTRODUCTION

Healthcare disparities, especially based on race and ethnicity, have been extensively documented across various medical conditions and stages of disease (1–3). Disparities manifest as unequal access to the healthcare system, differences in the quality of care, variations in medical device performance, and discrepancies in health outcomes not explained by clinical factors (4). They reflect systemic and societal biases that are easily incorporated into artificial intelligence (AI) models if researchers are not aware of them (5).

One of the main challenges in fair AI modeling is missingness handling (6,7). Notably, a 2017 study found that 49 out of 107 electronic health record (EHR)-based risk prediction approaches evaluated did not mention missing data at all (8). A common strategy is to replace missing values by physiologically-normal values. Similarly to clinical practice – where certain readings are only performed if there is a diagnostic suspicion – an “absent test” would be treated by the AI model as a “normal test” (9).

While this is considered a valid approach, it has important drawbacks. The likelihood of detecting an abnormal finding largely depends on how often a test is conducted. Thus, data that is not missing at random can drive spurious correlations, which are noncausal relationships between the input and the outcome which may shift in deployment (10). When the reason for missing data is a result of systemic social discrimination, these biases can be embedded in subsequent AI models, perpetuating and exacerbating existing disparities (11,12). Especially, as frequency of measurements has gained interest in the AI building community (13). As a case study of a potential source of such spurious correlations in medical AI models, we picked glucose measurement frequency among patients with sepsis admitted to the Intensive Care Unit (ICU).

Sepsis is a severe life-threatening systemic infection that has a significant impact on the health systems (14–16). Up to 90% of ICU patients exhibit elevated glucose levels irrespective of pre-existing diabetes (17–19). While elevated glucose has been linked to severity of illness, and increased mortality rates (20–22). Numerous studies have shown conflicting results, partly attributed to the detrimental effects of hypoglycemia, a consequence of strict glucose control (22–25). Sepsis patients who are admitted to the ICU are particularly vulnerable to blood glucose fluctuations (23,26) due to the inflammatory response and various aspects of care, such as corticosteroid use (27).

This paper aims to investigate potential disparities in glucose monitoring practices. We advocate that this kind of analysis must be done prior to any AI modeling. The primary objective is to determine if racial and ethnic or demographic differences influence the frequency of glucose measurements during sepsis management. By shedding light on this aspect of care, we aim to contribute to a more comprehensive understanding of healthcare inequalities and to provide researchers developing AI models with a framework to prevent potential biases adversely influencing model fairness and equity.

## METHODS

This observational retrospective study is reported in accordance with the Strengthening the Reporting of Observational Studies in Epidemiology (STROBE) statement (28). The health equity language, narrative and concepts of this paper follows the American Medical Association’s recommendations (29).

### Data Extraction

Data was extracted from the publicly-available MIMIC-IV database using SQL via Google’s BigQuery (30). The MIMIC database is maintained by the Laboratory for Computational Physiology at the Massachusetts Institute of Technology and shared via the PhysioNet platform (31). The dataset has been de-identified, and the institutional review boards of the Massachusetts Institute of Technology (No. 0403000206) and Beth Israel Deaconess Medical Center (2001-P-001699/14) both approved the use of the database for research. The MIMIC-IV database includes physiologic data collected from bedside monitors, laboratory test results, medications, medical images and clinical progress notes captured in the electronic health record from patients admitted to the ICU between 2008-2019. Approximately 70,000 de-identified medical records are archived in the MIMIC-IV database.

### Hypothesis

We hypothesized that the likelihood for a patient to receive measurement as well as the frequency of those measurements are not equally distributed across race and ethnicity, sex, or English proficiency.

### Cohort Selection

The following cases are excluded to create a study cohort: patients under 18 years of age, those without sepsis as defined by the sepsis 3 criteria (32), those with length of ICU stay less than 1 day, those with a diagnosis of diabetic ketoacidosis, and with racial description that does not fit within White, Asian, Black, or Hispanic, excluding those of the heterogenous group “other”.

### Covariates

We drew directed acyclic graphs to understand which variables to extract (see **Supplemental Figure 1 and 2**), as well as their interplay. A total of 13 patient-level variables were extracted, including non-time-varying variables such as demographics, comorbidities, and admission information and also time-varying variables including Sequential Organ Failure Assessment (SOFA) score (33), insulin treatment, glucocorticoid treatment, and instances of glucose measurements. Time-varying variables were aggregated in different ways. SOFA was calculated for the day of ICU admission, insulin treatment was used as a binary variable for whether or not it was received on day one. Glucose measurement was also used as a binary variable for whether or not it was measured on day one, in addition to taking the overall number of measurements for the whole ICU stay normalized by length of ICU stay. Glucocorticoid treatment was also normalized by ICU length of stay (see **Supplemental Table 1**).

### Outcomes

We had two primary outcomes: The first was a binary variable predicting whether a patient received measurement on day 1, and the second being a prediction of how many glucose measurements a patient would receive per day throughout the length of their ICU stay.

### Statistical Analysis

Statistical analysis was performed using Python 3.10. For summary statistics, we used the *tableone* Python package (34). For the outcome of whether or not a patient received a glucose measurement on day 1, we fitted a penalized, ridge logistic regression using the Python package *statsmodel* (35) adjusted for confounders to estimate the likelihood of receiving a glucose measurement. We report our findings as odds ratios (OR) with 95% confidence intervals (95% CI). White patients were considered as the reference group.

For the outcome of the number of glucose measurements during an ICU stay, we fitted a non-penalized, negative binomial regression (also with *statsmodel*) adjusted for confounders to estimate the number of glucose measurements for each patient each day during their respective ICU stay. We report our findings as incident rate ratios (IRR) with 95% CI. White patients were considered as the reference group.

## RESULTS

### Baseline Study Cohort

The MIMIC-IV database has 73,181 ICU stays, of which 24,927 (see **Figure 1**) made up our final cohort following the inclusion and exclusion criteria. The race and ethnicity distribution was 11.8% Black, 3.3% Asian, 3.3% Hispanic, 80.47% White and did not change relevantly after applying exclusion criteria.

**Figure 1.**
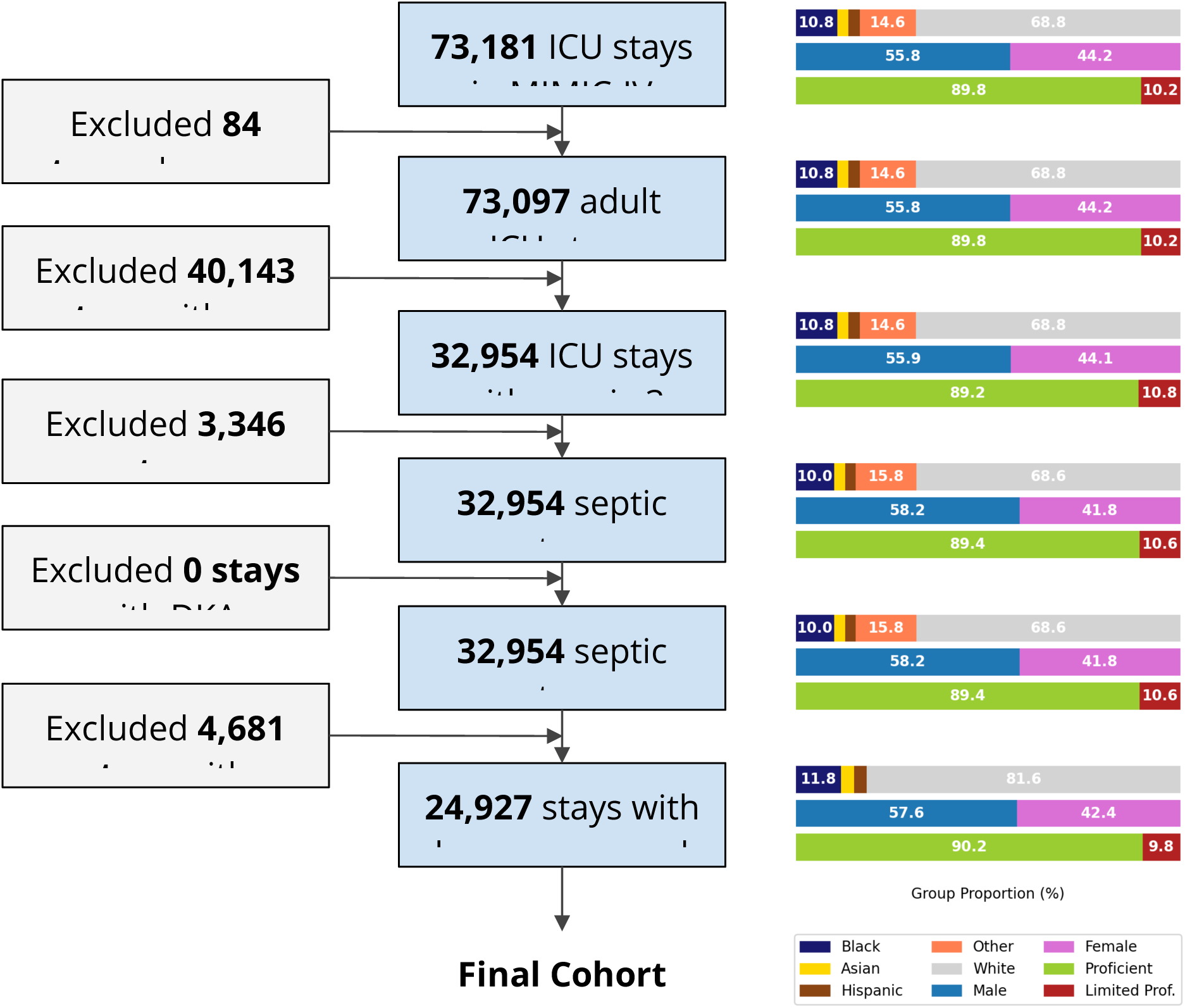
Consort diagram, depicting inclusion and exclusion criteria, as well as cohort composition at each step. DKA, Diabetic ketoacidosis; LOS, Length of stay

In the resulting cohort, English proficiency was distributed unevenly across race and ethnicity with Asian (37.8%) and Hispanic (39.3%) patients being significantly less frequently English proficient than White (95.2%) and Black (89.6%) patients (see **Table 1**). Insurance coverage also differed between race and ethnicity with White patients having the highest Medicare coverage (52.3%), and Hispanic patients had the highest Medicaid coverage (20.2%), followed by Asian patients (17.5%). Of note, even though the percentage of patients with diabetes was higher for Black (18.6%) and Hispanic (17.9%) patients when compared to White (11.4%) and Asian (14.2%) patients, the probability for receiving a glucose measurement on day 1 was roughly even for all groups (see **Supplementary Figure 3**). Furthermore, Hispanic (56.1%) patients had the lowest probability of receiving insulin on day 1.

**Table 1.**
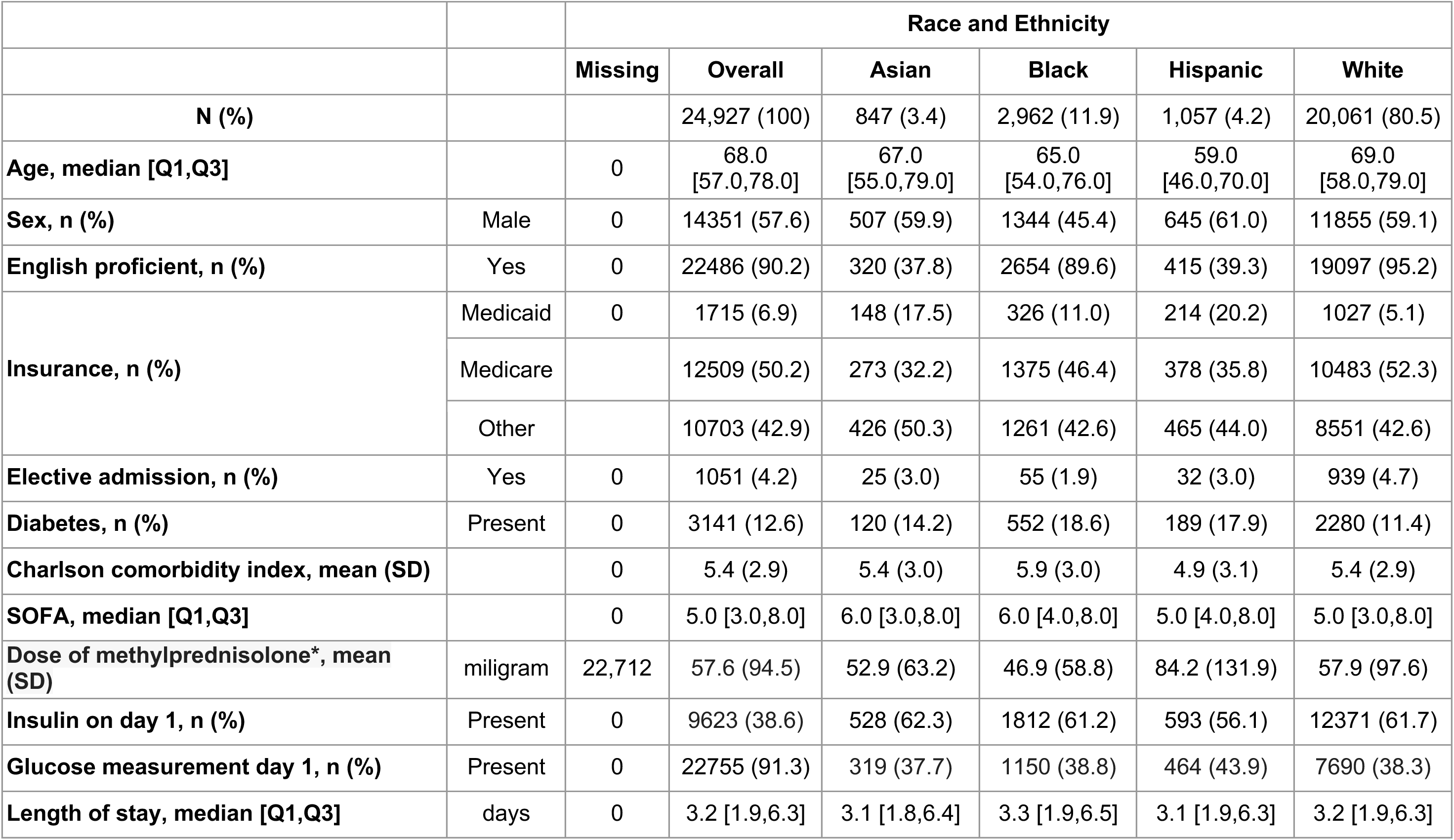
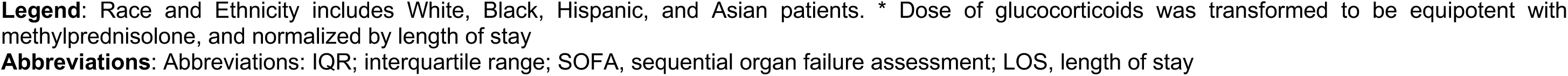
Baseline information on the study cohort, derived from MIMIC-IV.

|  |  | Race and Ethnicity |  |  |  |  |  |
| --- | --- | --- | --- | --- | --- | --- | --- |
|  |  | Missing | Overall | Asian | Black | Hispanic | White |
| **N (%)** |  |  | 24,927 (100) | 847 (3.4) | 2,962 (11.9) | 1,057 (4.2) | 20,061 (80.5) |
| **Age, median [Q1,Q3]** |  | 0 | 68.0<br>[57.0,78.0] | 67.0<br>[55.0,79.0] | 65.0<br>[54.0,76.0] | 59.0<br>[46.0,70.0] | 69.0<br>[58.0,79.0] |
| **Sex, n (%)** | Male | 0 | 14351 (57.6) | 507 (59.9) | 1344 (45.4) | 645 (61.0) | 11855 (59.1) |
| **English proficient, n (%)** | Yes | 0 | 22486 (90.2) | 320 (37.8) | 2654 (89.6) | 415 (39.3) | 19097 (95.2) |
| **Insurance, n (%)** | Medicaid | 0 | 1715 (6.9) | 148 (17.5) | 326 (11.0) | 214 (20.2) | 1027 (5.1) |
|  | Medicare |  | 12509 (50.2) | 273 (32.2) | 1375 (46.4) | 378 (35.8) | 10483 (52.3) |
|  | Other |  | 10703 (42.9) | 426 (50.3) | 1261 (42.6) | 465 (44.0) | 8551 (42.6) |
| **Elective admission, n (%)** | Yes | 0 | 1051 (4.2) | 25 (3.0) | 55 (1.9) | 32 (3.0) | 939 (4.7) |
| **Diabetes, n (%)** | Present | 0 | 3141 (12.6) | 120 (14.2) | 552 (18.6) | 189 (17.9) | 2280 (11.4) |
| **Charlson comorbidity index, mean (SD)** |  | 0 | 5.4 (2.9) | 5.4 (3.0) | 5.9 (3.0) | 4.9 (3.1) | 5.4 (2.9) |
| **SOFA, median [Q1,Q3]** |  | 0 | 5.0 [3.0,8.0] | 6.0 [3.0,8.0] | 6.0 [4.0,8.0] | 5.0 [4.0,8.0] | 5.0 [3.0,8.0] |
| **Dose of methylprednisolone\*, mean (SD)** | miligram | 22,712 | 57.6 (94.5) | 52.9 (63.2) | 46.9 (58.8) | 84.2 (131.9) | 57.9 (97.6) |
| **Insulin on day 1, n (%)** | Present | 0 | 9623 (38.6) | 528 (62.3) | 1812 (61.2) | 593 (56.1) | 12371 (61.7) |
| **Glucose measurement day 1, n (%)** | Present | 0 | 22755 (91.3) | 319 (37.7) | 1150 (38.8) | 464 (43.9) | 7690 (38.3) |
| **Length of stay, median [Q1,Q3]** | days | 0 | 3.2 [1.9,6.3] | 3.1 [1.8,6.4] | 3.3 [1.9,6.5] | 3.1 [1.9,6.3] | 3.2 [1.9,6.3] |

**Legend:** Race and Ethnicity includes White, Black, Hispanic, and Asian patients. \* Dose of glucocorticoids was transformed to be equipotent with methylprednisolone, and normalized by length of stay
**Abbreviations:** Abbreviations: IQR; interquartile range; SOFA, sequential organ failure assessment; LOS, length of stay

### Model Results

We used logistic regression to predict whether a patient received a glucose measurement on day 1 with the null hypothesis that all patients are equally likely to receive a measurement. Asian patients (OR 0.80, 95% CI 0.62 - 1.03), Black patients (OR 1.00, 95% CI 0.87 - 1.16), and Hispanic patients (OR 1.05, 95% CI 0.82 - 1.35) patients were compared to White patients as a reference (see **Figure 2A**, **Table 2, and Supplementary Figure 4**). In addition, the effect of being English proficient (OR 0.98, 95% CI 0.82 - 1.17) and being male (OR 0.94, 95% CI 0.82 - 1.17) were not statistically significant.

**Figure 2.**
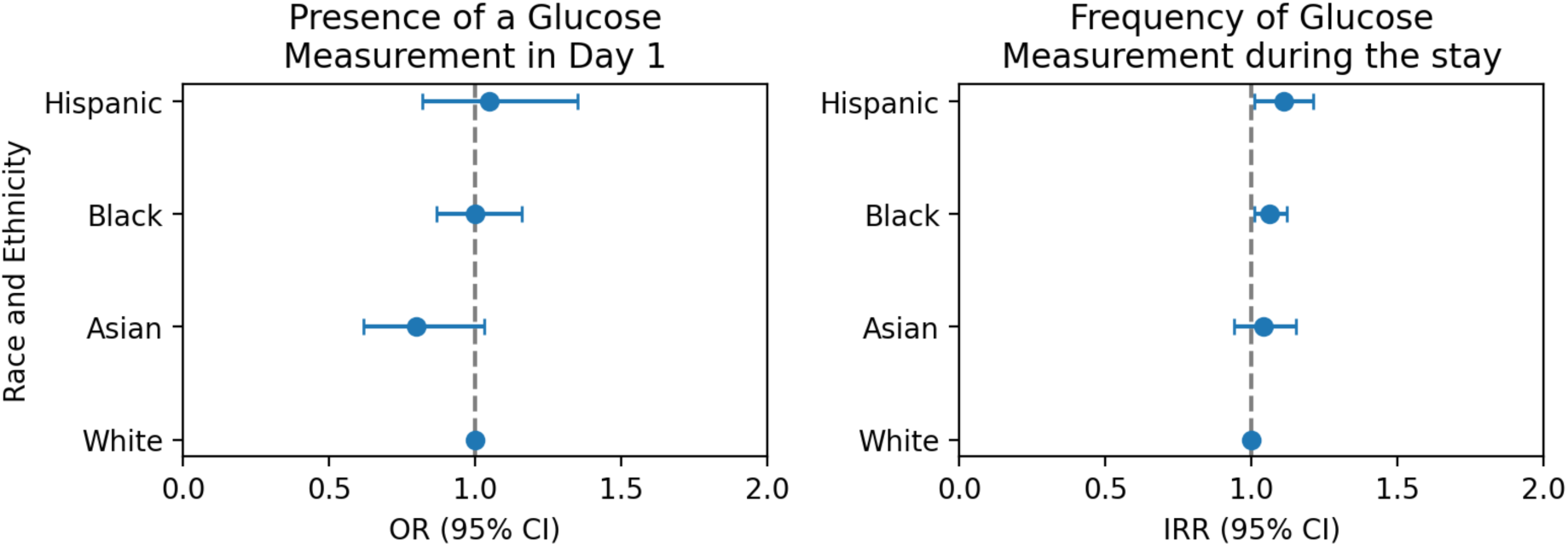
Main results, respective to race and ethnicity categorizations. Panel A: Results from logistic regression on measurement on day 1; Panel B: Results from negative binomial regression on frequency of measurements over whole ICU stay **Abbreviations**: OR, odds ratio; CI, confidence interval; IRR, incidence rate ratio

**Table 2.** Logistic regression results for outcome of glucose measurement on day 1.

| Variable | Odds Ratio | 2.5 % CI | 97.5 % CI |
| --- | --- | --- | --- |
| Intercept | 5.91 | 4.33 | 8.06 |
| Age | 1.00 | 1.00 | 1.00 |
| SOFA | 1.09 | 1.07 | 1.11 |
| Charlson comorbidity index | 1.05 | 1.03 | 1.07 |
| **Insurance:** |  |  |  |
| Other | Reference |  |  |
| Medicaid | 0.86 | 0.72 | 1.03 |
| Medicare | 0.97 | 0.87 | 1.07 |
| **Race and Ethnicity:** |  |  |  |
| White | Reference |  |  |
| Asian | 0.80 | 0.62 | 1.03 |
| Black | 1.00 | 0.87 | 1.16 |
| Hispanic | 1.05 | 0.82 | 1.35 |
| **Anchor year:** |  |  |  |
| 2008 - 2010 | Reference |  |  |
| 2011 - 2013 | 0.90 | 0.81 | 1.01 |
| 2014 - 2016 | 1.05 | 0.93 | 1.20 |
| 2017 - 2019 | 0.98 | 0.84 | 1.14 |
| **Binary variables:** |  |  |  |
| Male sex | 0.94 | 0.86 | 1.03 |
| Elective admission | 0.82 | 0.65 | 1.04 |
| English proficient | 0.98 | 0.82 | 1.17 |
| Major surgery | 1.92 | 1.73 | 2.14 |
| Glucocorticoids day one | 1.09 | 0.90 | 1.31 |
| English proficient | 0.98 | 0.82 | 1.17 |
| COPD present | 1.00 | 0.84 | 1.20 |
| Diabetes present | 1.72 | 1.43 | 2.05 |
| Asthma present | 1.38 | 0.55 | 3.47 |

Abbreviations: COPD, Chronic obstructive pulmonary disease; SOFA, sequential organ failure assessment

A negative binomial model was fitted to predict the total frequency of glucose measurements per patient and length of stay with the null hypothesis that all patients have the same frequency. Being male (IRR 1.06, 95% CI 1.01 - 1.21), Hispanic (IRR 1.11, 95% CI 1.01 - 1.21), Black (IRR 1.06, 95% CI 1.01 - 1.12), or English proficient (IRR 1.08, 95% CI 1.01 - 1.15) significantly increased the frequency of receiving a measurement (see **Figure 2B**, **Table 3, and Supplementary Figure 5**), while being Asian (IRR 1.04, 95% CI 0.94 - 1.15) did not.

**Table 3.** Negative binomial regression results for outcome of frequency of glucose measurement during length of stay.

| Variable | IRR | 2.5 % CI | 97.5 % CI |
| --- | --- | --- | --- |
| Intercept | 3.17 | 2.82 | 3.57 |
| Age | 1.00 | 1.00 | 1.00 |
| SOFA | 1.03 | 1.02 | 1.03 |
| Charlson comorbidity index | 0.99 | 0.99 | 1.00 |
| Methylprednisolone dose normalized by LOS | 1.00 | 1.00 | 1.00 |
| **Insurance:** |  |  |  |
| Other | Reference |  |  |
| Medicare | 0.98 | 0.94 | 1.02 |
| Medicaid | 0.94 | 0.88 | 1.01 |
| **Race:** |  |  |  |
| White | Reference |  |  |
| Asian | 1.04 | 0.94 | 1.15 |
| Black | 1.06 | 1.01 | 1.12 |
| Hispanic | 1.11 | 1.01 | 1.21 |
| **Anchor year:** |  |  |  |
| 2008 - 2010 | Reference |  |  |
| 2011 - 2013 | 0.96 | 0.92 | 1.00 |
| 2014 - 2016 | 1.01 | 0.96 | 1.05 |
| 2017 - 2019 | 0.93 | 0.88 | 0.98 |
| **Binary variables:** |  |  |  |
| Male sex | 1.06 | 1.03 | 1.10 |
| Elective admission | 1.46 | 1.34 | 1.58 |
| Insulin on day 1 | 1.69 | 1.64 | 1.75 |
| English proficient | 1.08 | 1.01 | 1.15 |
| Major surgery | 1.57 | 1.51 | 1.63 |
| COPD present | 0.93 | 0.87 | 0.99 |
| Diabetes present | 1.36 | 1.29 | 1.43 |
| Asthma present | 0.94 | 0.68 | 1.29 |

**Abbreviations:** IRR, incidence rate ratio; COPD, chronic obstructive pulmonary disease; SOFA, sequential organ failure assessment; LoS, length of stay

## DISCUSSION

Policymakers have increasingly recognized the importance of health equity to public health(36). In December of 2022, tying reimbursements to health equity outcomes was proposed by the Center for Medicare and Medicaid Services (CMS)(37). If these policies were to be implemented, they would also need to be considered in AI development as disparities could have unintended consequences on a model’s prediction (38). There is growing awareness in the domain of medical AI development that potential advancements through AI could be hindered by such biases (39). The low accuracy of the EPIC sepsis model in external datasets is a good example of what can happen if temporal bias is neglected(40). Another study found increased illness burden in Black patients because the model wrongly included the post-treatment cost as a pre-treatment feature(41).

Aside from the risks of inputting biased data into a model, unmeasured data for a particular group reflect quality of care. Absence of data in medicine is not simply a void. In fact, caregivers often do not measure a variable deliberately; the reason to do so is usually highly confounded, affected by ongoing treatment, and reflected in the potential outcome. Models also interpret these voids as information influencing predictions, driving spurious correlations (10). Therefore, it is important to establish a thorough understanding of such associations before fitting any AI model.

To avoid encrypting biases into AI models, many advocate the use of causal inference frameworks to check for, understand, and control for potential biases (42,43). Performing similar analyses as we did – even before fitting a causal inference model – should become a common practice, as the awareness and understanding of such incongruences can help to understand these models better.

Furthermore, one key assumption of causal inference models is positivity, i.e. subjects from the treatment and control group should have enough overlap in the distribution of their confounding variables(44). Usually researchers check this assumption by tabulating the data across the most pertinent variables which can become unwieldy in case of many strata. As an additional check, we propose fitting simple regression models as we did to ascertain that the positivity assumption is not being violated and adding frequency of measurements as a distinct feature to any model.

Besides all these implications for AI models, our findings are also relevant clinically. While our study found no differences on glucose measurements in the glucose measurements on day 1 among patients admitted to the ICU due to sepsis, there was a slightly higher frequency of glucose measurement among Black, Hispanic, male, and English proficient patients.

Glucose measurement plays a pivotal role in in-patient care, as glucose levels are linked to severity of the disease and patient outcomes (20,21). In the context of sepsis patients, hyperglycemia is a common bystander associated with severity of disease and mortality (23). Studies have shown that frequent glucose measurements are associated with less hypoglycemia and hyperglycemia episodes, as well as decreased glucose variability (45–48) which has been associated with decreased hospital mortality, decreased length of hospital stay, and composite hospital complications (48). Reports of Black patients admitted to the ICU having poorer glycemic control are thus troublesome (49).

## LIMITATIONS

While our study adds to the discussion of disparities and algorithmic bias in the critical care setting, we acknowledge that it has certain limitations. These include potential selection bias, as the database does not include patients who were admitted to the regular ward, and the data comes from only one academic tertiary care center in the U.S, where most of the population is White (80.47%). Also, our study design does not allow us to test whether the associations are from unmeasured confounding. Future research should strive to address these limitations for a more comprehensive understanding of the topic and should also cover other areas of care such as emergency departments, regular wards, or ambulatory care.

## CONCLUSION

The implications of our study goes beyond glucose monitoring during sepsis management. Besides the challenge of achieving healthcare equity in a system marked by systemic biases, researchers need to be aware of such disparities before building any AI model. Such biases cannot only skew predictions, but may even generate further adverse effects when the predictions are being used for treatment or management decisions.

## Author Contributions

All authors contributed to writing the manuscript.

KT and YJ performed the statistical analysis.

NLW wrote the first draft of the manuscript.

LAC, JM, and TS provided idea, concept, technical and administrative support.

## Data Availability

The data that support the findings of this study are available in MIMIC-IV with the identifier doi.org/10.1093/jamia/ocx084 publicly available on PhysioNet (https://physionet.org/).

## Code Availability

The code that produces the results in this manuscript can be accessed at https://github.com/e754/Glucose-Research, which includes detailed instructions for running the code.

## SUPPLEMENTARY MATERIALS

**Supplementary Figure 1.**
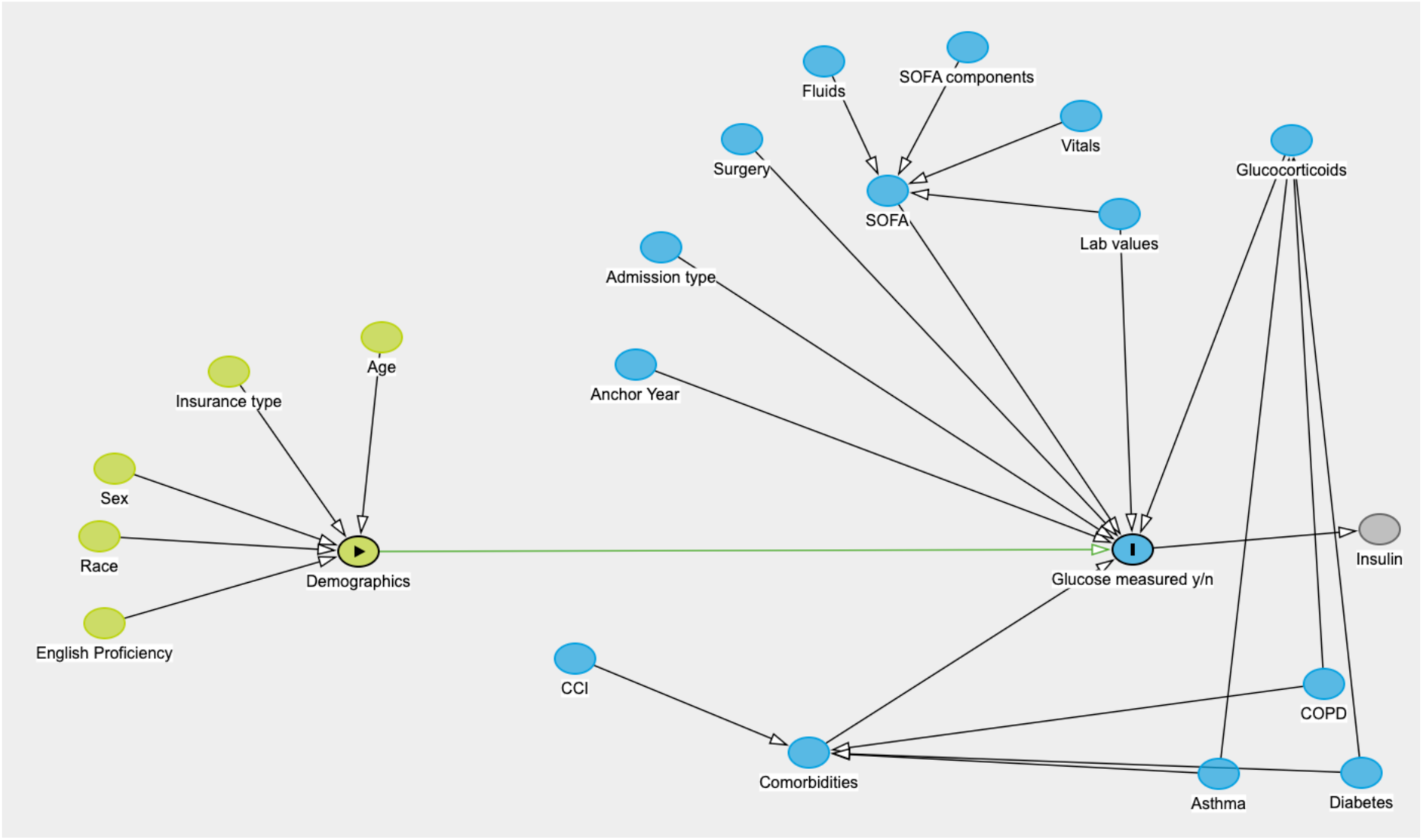
Directed acyclic graph for outcome of a single glucose measurement on day 1 of hospital admission. Race includes White, Black, Hispanic, and Asian patients. **Abbreviations**: IQR; interquartile range; COPD, chronic obstructive pulmonary disease; SOFA, sequential organ failure assessment; CCI, Charlson Comorbidity Index. Lab, laboratory

**Supplementary Figure 2.**
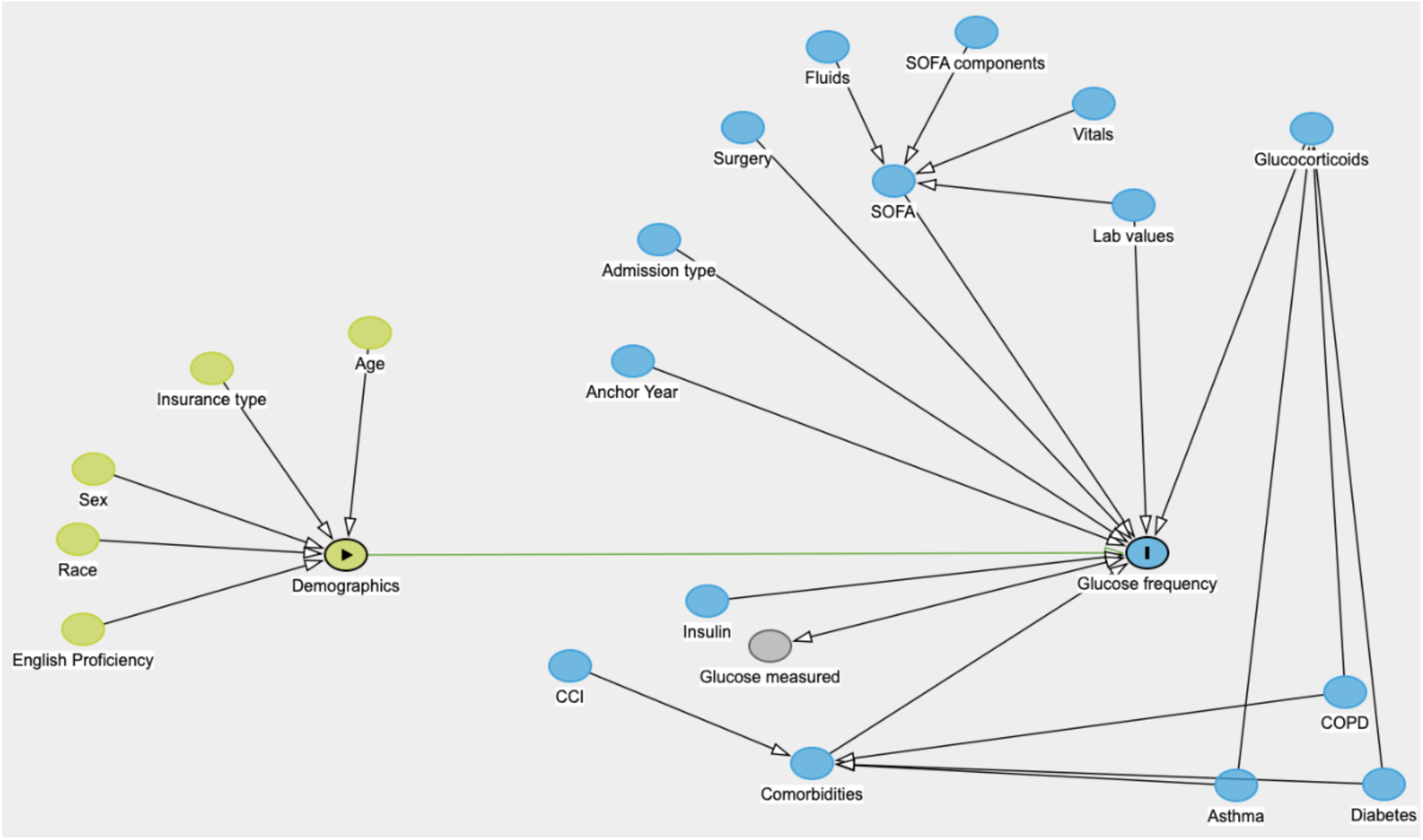
Directed acyclic graph for outcome of frequency of glucose measurement over the hospital admission. Race includes White, Black, Hispanic, and Asian patients. **Abbreviations**: IQR; interquartile range; COPD, chronic obstructive pulmonary disease; SOFA, sequential organ failure assessment; CCI, Charlson Comorbidity Index. Lab, laboratory

**Supplementary Figure 3.**
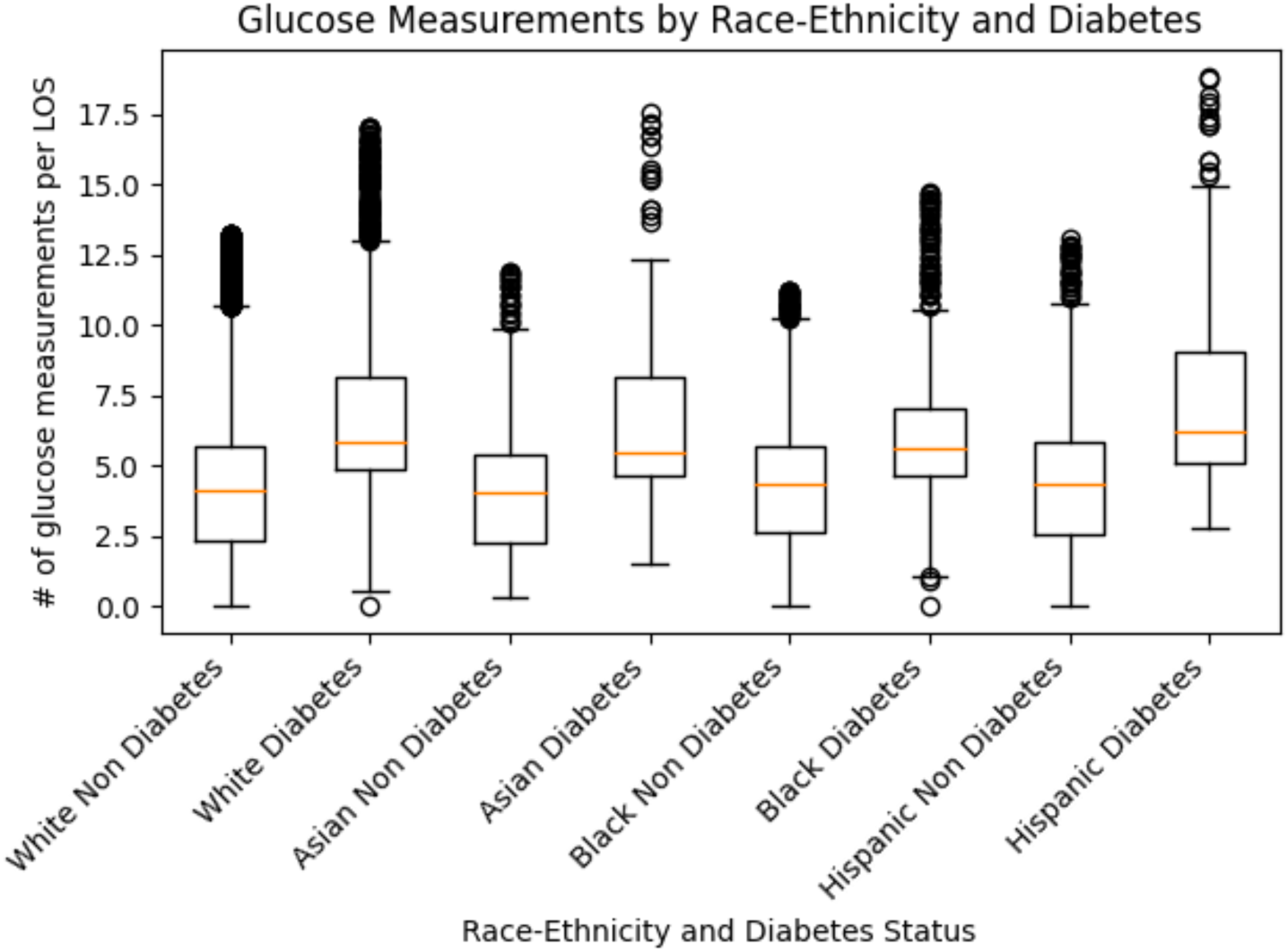
Boxplot of number of glucose measurements per LOS binned by race-ethnicity and stratified by diabetes status. Race includes White, Black, Hispanic, and Asian patients. Orange horizontal bar denotes the median, while the box encompasses the first quartile to the third quartile. The whiskers denote 1.5x the interquartile range. Points are outliers. **Abbreviations**: #, number; LOS, length of ICU stay.

**Supplementary Figure 4.**
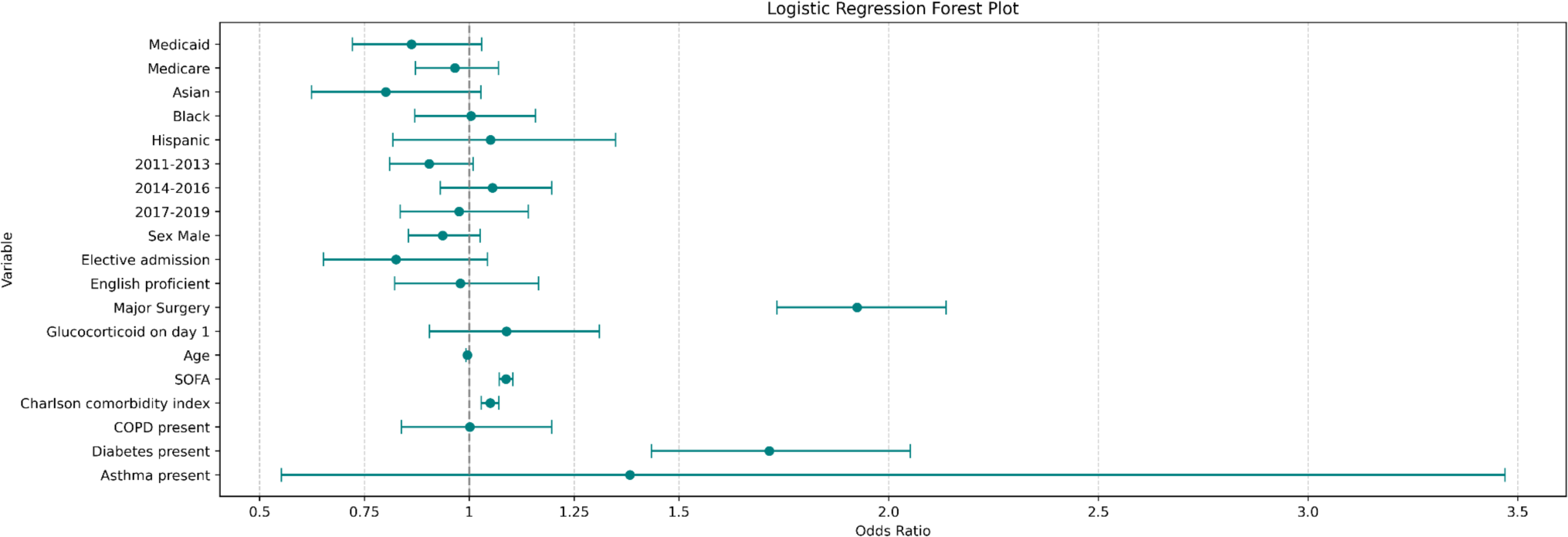
Logistic Regression model forest plot.

**Supplementary Figure 5.**
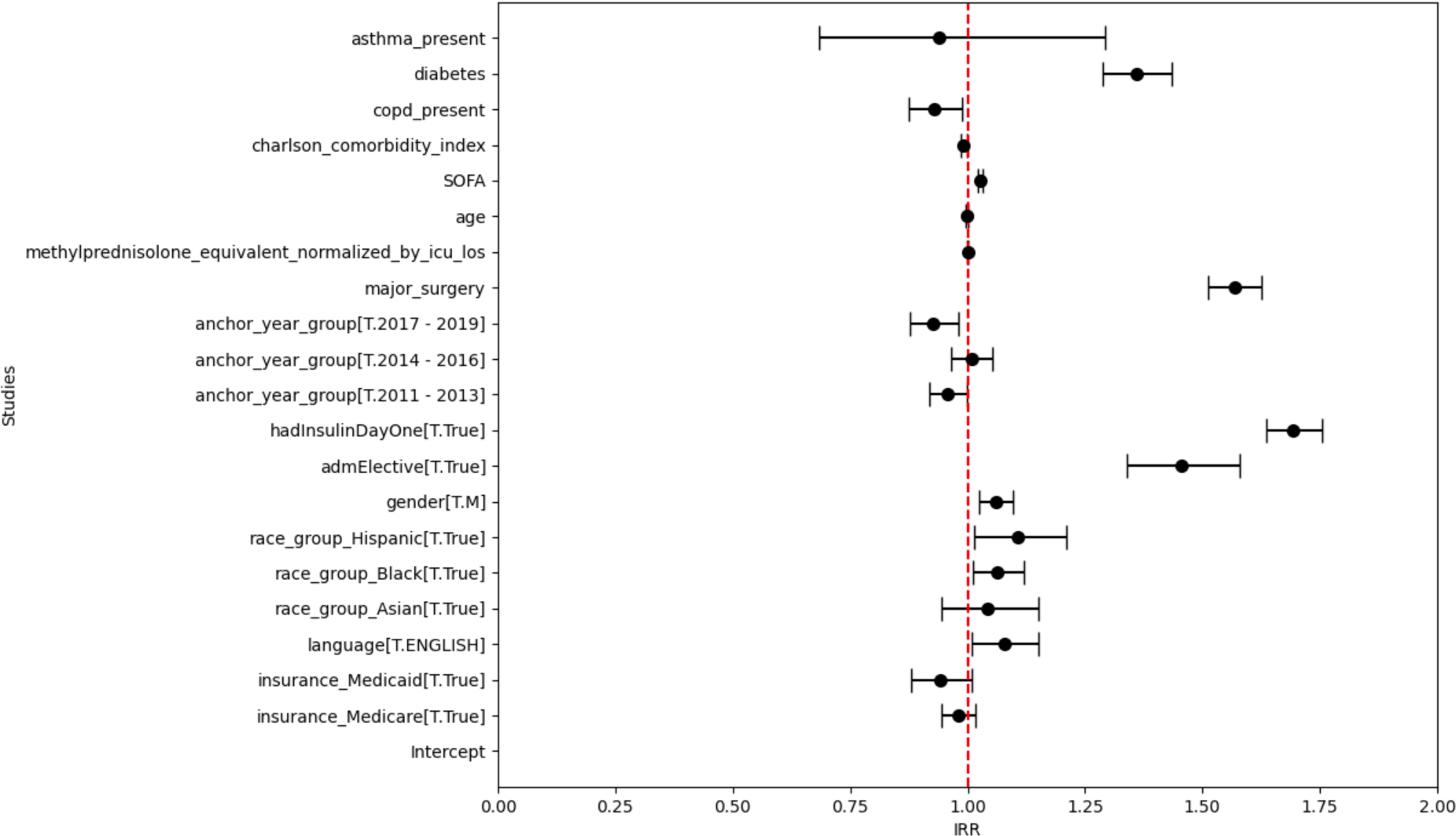
Negative Binomial Model Forest Plot.

**Supplementary Table 1.** Definition of considered covariates.

| **Covariates** | **Description** | **Handling of missing values** |
| --- | --- | --- |
| Sex | As provided by dataset | N/A |
| Age | At admission | N/A |
| Race Ethnicity | Grouped into Asian, Black, White, Hispanic, and Other | Exclusion of group "other" |
| Insurance | As provided by dataset, Medicare/Medicaid or other | N/A |
| English Proficiency | As provided by dataset, English or limited | N/A |
| Year Group | As provided by dataset, bi-yearly bins | N/A |
| Elective Admission | As provided by dataset, true or false if admission is elective | N/A |
| Major Surgery | As provided by OASIS score | N/A |
| SOFA | SOFA score with each of its subcomponents: on admission and for the selected 24 hours, aggregated by the maximum value | Assumption of best possible value in case of missing |
| Charlson comorbidity Index | As provided by dataset | N/A |
| COPD | ICD-10 codes J41.X-J47.X | N/A |
| Asthma | ICD-10 codes J84.1X | N/A |
| Diabetes | ICD-10 codes E08.X-E11.X, and E13.X<br>ICD-10 codes L94.0X, L94.1X, L94.3X | N/A |
| DKA present | ICD-10 codes E10.1X, E11.1X, E13.1X | N/A |
| Insulin | MIMIC-IV Item IDs 229299, 229619, 223257, 223258, 223259, 223260, 223261, 223262 | Assume no insulin received if no entries. |
| Glucose | MIMIC-IV Item IDs 220621, 228388, 25664, 226537 | Assume no glucose measurement if no entries. |
| Methylprednisolone dose normalized by LOS | Only non-topical formulations of Dexamethasone, Hydrocortisone, Methylprednisolone, and Prednisone were considered.<br>Methylprednisolone equivalent doses were calculated by multiplying the dose with following factors:<br>Dexamethasone x5<br>Hydrocortisone x0.2<br>Methylprednisolone x1<br>Prednisone x1 | Assume no glucocorticoid received if no entries. |

## REFERENCES

1. Hall WJ, Chapman MV, Lee KM, Merino YM, Thomas TW, Payne BK, et al. Implicit Racial/Ethnic Bias Among Health Care Professionals and Its Influence on Health Care Outcomes: A Systematic Review. Am J Public Health. 2015 Dec;105(12):e60–76.

2. Magesh S, John D, Li WT, Li Y, Mattingly-App A, Jain S, et al. Disparities in COVID-19 Outcomes by Race, Ethnicity, and Socioeconomic Status: A Systematic-Review and Meta-analysis. JAMA Netw Open. 2021 Nov 1;4(11):e2134147.

3. Vela MB, Erondu AI, Smith NA, Peek ME, Woodruff JN, Chin MH. Eliminating Explicit and Implicit Biases in Health Care: Evidence and Research Needs. Annu Rev Public Health. 2022 Apr 5;43:477–501.

4. Charpignon ML, Byers J, Cabral S, Celi LA, Fernandes C, Gallifant J, et al. Critical Bias in Critical Care Devices. Crit Care Clin. 2023 Oct 1;39(4):795–813.

5. Ferryman K, Mackintosh M, Ghassemi M. Considering Biased Data as Informative Artifacts in AI-Assisted Health Care. N Engl J Med. 2023 Aug 31;389(9):833–8.

6. Chen IY, Pierson E, Rose S, Joshi S, Ferryman K, Ghassemi M. Ethical Machine Learning in Healthcare. Annu Rev Biomed Data Sci. 2021;4(1):123–44.

7. Nazer LH, Zatarah R, Waldrip S, Ke JXC, Moukheiber M, Khanna AK, et al. Bias in artificial intelligence algorithms and recommendations for mitigation. PLOS Digit Health. 2023 Jun 22;2(6):e0000278.

8. Gold R, Cottrell E, Bunce A, Middendorf M, Hollombe C, Cowburn S, et al. Developing Electronic Health Record (EHR) Strategies Related to Health Center Patients’ Social Determinants of Health. J Am Board Fam Med. 2017 Jul;30(4):428–47.

9. Wells BJ, Nowacki AS, Chagin K, Kattan MW. Strategies for Handling Missing Data in Electronic Health Record Derived Data. 2013 Dec 17;1(3):7.

10. Yang Y, Zhang H, Katabi D, Ghassemi M. Change is Hard: A Closer Look at Subpopulation Shift [Internet]. arXiv; 2023 [cited 2023 Sep 27]. Available from: http://arxiv.org/abs/2302.12254

11. Panch T, Mattie H, Atun R. Artificial intelligence and algorithmic bias: implications for health systems. J Glob Health. 9(2):020318.

12. Mattu JA Jeff Larson, Lauren Kirchner, Surya. ProPublica. [cited 2023 Sep 29]. Machine Bias. Available from: https://www.propublica.org/article/machine-bias-risk-assessments-in-criminal-sentencing

13. Chang CH, Mai M, Goldenberg A. Dynamic Measurement Scheduling for Event Forecasting using Deep RL. In: Proceedings of the 36th International Conference on Machine Learning [Internet]. PMLR; 2019 [cited 2023 Sep 30]. p. 951–60. Available from: https://proceedings.mlr.press/v97/chang19a.html

14. Rhee C, Dantes R, Epstein L, Murphy DJ, Seymour CW, Iwashyna TJ, et al. Incidence and Trends of Sepsis in US Hospitals Using Clinical vs Claims Data, 2009-2014. JAMA. 2017 Oct 3;318(13):1241–9.

15. Seymour CW, Gesten F, Prescott HC, Friedrich ME, Iwashyna TJ, Phillips GS, et al. Time to Treatment and Mortality during Mandated Emergency Care for Sepsis. N Engl J Med. 2017 Jun 8;376(23):2235–44.

16. Singer M, Deutschman CS, Seymour CW, Shankar-Hari M, Annane D, Bauer M, et al. The Third International Consensus Definitions for Sepsis and Septic Shock (Sepsis-3). JAMA. 2016 Feb 23;315(8):801–10.

17. Levetan CS, Passaro M, Jablonski K, Kass M, Ratner RE. Unrecognized diabetes among hospitalized patients. Diabetes Care. 1998 Feb;21(2):246–9.

18. Umpierrez GE, Isaacs SD, Bazargan N, You X, Thaler LM, Kitabchi AE. Hyperglycemia: an independent marker of in-hospital mortality in patients with undiagnosed diabetes. J Clin Endocrinol Metab. 2002 Mar;87(3):978–82.

19. Zhu W, Jiang L, Jiang S, Ma Y, Zhang M. Real-time continuous glucose monitoring versus conventional glucose monitoring in critically ill patients: a systematic review study protocol. BMJ Open. 2015 Jan 23;5(1):e006579.

20. ACE/ADA Task Force on Inpatient Diabetes. American College of Endocrinology and American Diabetes Association Consensus statement on inpatient diabetes and glycemic control. Diabetes Care. 2006 Aug;29(8):1955–62.

21. Yamada T, Shojima N, Noma H, Yamauchi T, Kadowaki T. Glycemic control, mortality, and hypoglycemia in critically ill patients: a systematic review and network meta-analysis of randomized controlled trials. Intensive Care Med. 2017 Jan;43(1):1–15.

22. van den Berghe G, Wouters P, Weekers F, Verwaest C, Bruyninckx F, Schetz M, et al. Intensive insulin therapy in critically ill patients. N Engl J Med. 2001 Nov 8;345(19):1359–67.

23. Silveira LM, Basile-Filho A, Nicolini EA, Dessotte C a. M, Aguiar GCS, Stabile AM. Glycaemic variability in patients with severe sepsis or septic shock admitted to an Intensive Care Unit. Intensive Crit Care Nurs. 2017 Aug;41:98–103.

24. Uyttendaele V, Dickson JL, Shaw GM, Desaive T, Chase JG. Untangling glycaemia and mortality in critical care. Crit Care Lond Engl. 2017 Jun 24;21(1):152.

25. Yatabe T, Inoue S, Sakaguchi M, Egi M. The optimal target for acute glycemic control in critically ill patients: a network meta-analysis. Intensive Care Med. 2017 Jan;43(1):16– 28.

26. Ellger B, Westphal M, Stubbe HD, Van den Heuvel I, Van Aken H, Van den Berghe G. [Glycemic control in sepsis and septic shock: friend or foe?]. Anaesthesist. 2008 Jan;57(1):43–8.

27. Andersen SK, Gjedsted J, Christiansen C, Tønnesen E. The roles of insulin and hyperglycemia in sepsis pathogenesis. J Leukoc Biol. 2004 Mar;75(3):413–21.

28. Cuschieri S. The STROBE guidelines. Saudi J Anaesth. 2019 Apr;13(Suppl 1):S31–4.

29. Flanagin A, Frey T, Christiansen SL, AMA Manual of Style Committee. Updated Guidance on the Reporting of Race and Ethnicity in Medical and Science Journals. JAMA. 2021 Aug 17;326(7):621–7.

30. Johnson AEW, Bulgarelli L, Shen L, Gayles A, Shammout A, Horng S, et al. MIMIC-IV, a freely accessible electronic health record dataset. Sci Data. 2023 Jan 3;10(1):1.

31. Goldberger AL, Amaral LA, Glass L, Hausdorff JM, Ivanov PC, Mark RG, et al. PhysioBank, PhysioToolkit, and PhysioNet: components of a new research resource for complex physiologic signals. Circulation. 2000 Jun 13;101(23):E215–220.

32. Singer M, Deutschman CS, Seymour CW, Shankar-Hari M, Annane D, Bauer M, et al. The Third International Consensus Definitions for Sepsis and Septic Shock (Sepsis-3). JAMA. 2016 Feb 23;315(8):801–10.

33. Vincent JL, Moreno R, Takala J, Willatts S, De Mendonça A, Bruining H, et al. The SOFA (Sepsis-related Organ Failure Assessment) score to describe organ dysfunction/failure. On behalf of the Working Group on Sepsis-Related Problems of the European Society of Intensive Care Medicine. Intensive Care Med. 1996 Jul;22(7):707– 10.

34. Pollard TJ, Johnson AEW, Raffa JD, Mark RG. tableone: An open source Python package for producing summary statistics for research papers. JAMIA Open. 2018 May 23;1(1):26–31.

35. Seabold S, Perktold J. statsmodels: Econometric and statistical modeling with python. In: 9th Python in Science Conference. 2010.

36. Health Equity and Health Disparities Environmental Scan. FINAL Rep.

37. CMS Proposes Policies to Improve Patient Safety and Promote Health Equity | CMS [Internet]. [cited 2023 Sep 26]. Available from: https://www.cms.gov/newsroom/press-releases/cms-proposes-policies-improve-patient-safety-and-promote-health-equity

38. Celi LA, Cellini J, Charpignon ML, Dee EC, Dernoncourt F, Eber R, et al. Sources of bias in artificial intelligence that perpetuate healthcare disparities—A global review. PLOS Digit Health. 2022 Mar 31;1(3):e0000022.

39. Saint James Aquino Y. Making decisions: Bias in artificial intelligence and data-driven diagnostic tools. Aust J Gen Pract. 2023 Jul;52(7):439–42.

40. Wong A, Otles E, Donnelly JP, Krumm A, McCullough J, DeTroyer-Cooley O, et al. External Validation of a Widely Implemented Proprietary Sepsis Prediction Model in Hospitalized Patients. JAMA Intern Med. 2021 Aug 1;181(8):1065–70.

41. Obermeyer Z, Powers B, Vogeli C, Mullainathan S. Dissecting racial bias in an algorithm used to manage the health of populations. Science. 2019 Oct 25;366(6464):447–53.

42. Schölkopf B, Locatello F, Bauer S, Ke NR, Kalchbrenner N, Goyal A, et al. Toward Causal Representation Learning. Proc IEEE. 2021 May;109(5):612–34.

43. Plecko D, Bareinboim E. Causal Fairness Analysis [Internet]. arXiv; 2022 [cited 2023 Sep 28]. Available from: http://arxiv.org/abs/2207.11385

44. Doutreligne M, Struja T, Abecassis J, Morgand C, Celi LA, Varoquaux G. Causal thinking for decision making on Electronic Health Records: why and how [Internet]. arXiv; 2023 [cited 2023 Sep 28]. Available from: http://arxiv.org/abs/2308.01605

45. Krinsley JS, Bruns DE, Boyd JC. The Impact of Measurement Frequency on the Domains of Glycemic Control in the Critically Ill-A Monte Carlo Simulation. J Diabetes Sci Technol. 2015 Mar 1;9(2):237–45.

46. Jacobi J, Bircher N, Krinsley J, Agus M, Braithwaite SS, Deutschman C, et al. Guidelines for the use of an insulin infusion for the management of hyperglycemia in critically ill patients. Crit Care Med. 2012 Dec;40(12):3251–76.

47. Sreedharan R, Martini A, Das G, Aftab N, Khanna S, Ruetzler K. Clinical challenges of glycemic control in the intensive care unit: A narrative review. World J Clin Cases. 2022 Nov 6;10(31):11260–72.

48. Falciglia M, Freyberg RW, Almenoff PL, D’Alessio DA, Render ML. Hyperglycemia-related mortality in critically ill patients varies with admission diagnosis. Crit Care Med. 2009 Dec;37(12):3001–9.

49. Fayfman M, Vellanki P, Alexopoulos AS, Buehler L, Zhao L, Smiley D, et al. Report on Racial Disparities in Hospitalized Patients with Hyperglycemia and Diabetes. J Clin Endocrinol Metab. 2016 Mar;101(3):1144–50.

